# The impact of depression and childhood maltreatment experiences on psychological adaptation from lockdown to relaxation periods during the COVID-19 pandemic

**DOI:** 10.1101/2023.10.10.23296796

**Authors:** Julian Herpertz, Janik Goltermann, Marius Gruber, Rogério Blitz, Jacob Taylor, Katharina Brosch, Frederike Stein, Benjamin Straube, Susanne Meinert, Anna Kraus, Elisabeth J Leehr, Jonathan Repple, Ronny Redlich, Lara Gutfleisch, Bianca Besteher, Janette Ratzsch, Alexandra Winter, Linda M. Bonnekoh, Daniel Emden, Tilo Kircher, Igor Nenadic, Udo Dannlowski, Tim Hahn, Nils Opel

## Abstract

The COVID-19 pandemic has presented a significant challenge to societal mental health. Yet, it remains unknown which factors influence the mental adaptation from lockdown to subsequent relaxation periods, particularly for vulnerable groups.

This study used smartphone-based monitoring to explore how 74 individuals with major depression (MDD) and 77 healthy controls (HCs) responded to the transition from lockdown to relaxation during the first wave of the COVID-19 pandemic (March 21 to November 01, 2020) regarding interpersonal interactions, COVID-19-related fear (fear of participants’ own health, the health of close relatives, and the pandemics’ economic impact), and the feeling of isolation. Furthermore, we investigated the effect of a diagnosis of MDD and the experience of childhood maltreatment (CM) on adaptive functioning.

During the transition from lockdown to relaxation, we observed an increase in direct contacts and a decrease in indirect contacts and self-perceived isolation in the study population. The diagnosis of MDD and the experience of CM moderated a maintenance of COVID-19-related fear: HCs and participants without the experience of CM showed a decrease in fear, while fear of participants with MDD and with an experience of CM did not change significantly.

The finding that elevated COVID-19-related fear was sustained in vulnerable groups after lockdown measures were lifted could help guide psychosocial prevention efforts in future pandemic emergencies.

## 1. Introduction

In 2019, the coronavirus pandemic (COVID-19) was caused by the outbreak and global spread of the SARS-CoV-2 virus. In response, governments took action to contain the spread of the virus. Individual behaviors were restricted to limit transmission rates. Restrictions in Germany included various forms of social distancing, limiting direct social contacts, closing schools and restaurants, and, in some states, requiring people to stay home. While restrictions reduced transmission rates in most countries (Chung et al., 2021), they inevitably led to unintended negative side effects, as the pandemic was associated with an increase in social anxiety, the isolation of segments of the population, and other insecurities that place an unprecedented psychological burden on society (Clair et al., 2021; Leung et al., 2022; Sojli et al., 2021).

It is important to note that, based on our current understanding, the burden of the pandemic on mental health was not homogeneously distributed among the general population but was more pronounced in populations with increased vulnerability for mental health problems (Ahrens et al., 2021a). Due to a greater susceptibility to stress and a higher likelihood of contracting the virus compared to the general population, it is assumed that many patients with a psychiatric disorder have experienced relapse or symptom exacerbation during the COVID-19 pandemic (Gobbi et al., 2020; Yao et al., 2020). Favreau and colleagues (2021) found that out of 500 inpatients with various mental health disorders, almost half of the patients experienced a worsening of symptoms during the first wave of COVID-19 in Germany (April 01 to December 31, 2020), with individuals facing depression being notably susceptible to symptom exacerbation. This prevalence exceeds rates of mental health deterioration in the general population (Xiong et al., 2020). In particular, contact restrictions and the obligation to stay at home were perceived as burdensome by the study population that was investigated by Favreau and colleagues. In another study conducted by Hao and colleagues (2020), it was found that patients with affective or anxiety disorders were significantly more likely to develop general and COVID-19-related anxiety during the pandemic than healthy controls (HCs).

Childhood maltreatment (CM) is a major risk factor for the development and an unfavorable course of disease in major depressive disorder (MDD; Opel et al., 2019). Moreover, the experience of CM has also been shown to predict the experience of severe stress from the pandemic (Medeiros et al., 2020; Rek et al., 2022). In a study investigating the relationship between CM and state anxiety during the COVID-19 pandemic, Kalia and colleagues (2020) found that subjects who experienced CM were not only more likely to perceive COVID-19 as an environmental threat, but this perception also mediated the relationship between maltreatment and state anxiety. Siegel and Lahav (2022) identified an association between CM and more overall COVID-19-related psychological distress and peritraumatic stress symptoms than in the control group. Previous exposure to traumatic events has also been identified as a predictor of the feeling of loneliness during the pandemic (Thakur et al., 2023).

While aforementioned studies investigated mainly the impact of governmental restrictions, the adaptation to relaxation periods following long periods of restrictions is widely understudied. However, relaxation succeeding restrictions also poses an adaptive challenge which could potentially impact mental health outcomes. Given the above findings on the influence of psychiatric diagnoses and CM on mental health during the pandemic, it appears specifically relevant to investigate these influences also with respect to such adaptation processes.

By means of longitudinal smartphone-based monitoring the present study investigated how participants adapted to the change from lockdown to relaxation in Germany throughout 2020. Furthermore, we investigated how the diagnosis of MDD and the experience of CM was associated with a dysfunctional adaptation in comparison to the control group. Aforementioned research highlights the worsening of mental well-being within vulnerable groups during the pandemic, especially in relation to the constraints on interpersonal interactions, COVID-19-related fear (fear in relation to participants’ own health, the health of close relatives, and the pandemics’ economic impact), and the feeling of isolation associated with the pandemic. Consequently, our study used social behavior encompassing both direct and indirect contacts, COVID-19-related fear, and feelings of isolation as parameters governing mental health outcomes. We hypothesize:

1. an increase of direct contacts and a decrease of indirect contacts, a decrease of COVID-19-related fear and reduced feelings of isolation for the whole study sample during the transition from lockdown to relaxation.
2. an attenuated change in social behavior, COVID-19-related fear, and feelings of isolation in subjects with previous diagnosis of MDD compared to HCs and in subjects with experience of CM compared to subjects without the experience of CM.

## 2. Methods

### 2.1 Smartphone-based data acquisition: *ReMAP*

ReMAP (Remote Monitoring Application for Psychiatry) was implemented at the Institute for Translational Psychiatry in Muenster, Germany in 2018 (Emden et al., 2021; Goltermann et al., 2021). It was designed as a complementary evaluation method for ongoing longitudinal observational studies. The data collected serve as an add-on assessment to previously evaluated data units from other studies, e.g., Magnetic Resonance Imaging (MRI) data, genetics and microbiome data, neuropsychological tests, clinical interviews, and a variety of clinical and personality questionnaires. ReMAP is a native app for iOS and Android, based on Apple ResearchKit, Apple Health, and Google Fit. Once installed, the app works in the background of the phone and monitors the number of steps and the covered distance during a day. Additionally, it regularly asks participants to give feedback on their mood, their depressive symptoms (via the Beck’s Depression Inventory; BDI; Beck et al., 1960) and on their sleep through self-report questionnaires. The ReMAP app, its protocols and data entities were comprehensively described, and the validity and feasibility has been shown in previous studies (Emden et al., 2021; Goltermann et al., 2021). During the first lockdown of the COVID-19 pandemic in Germany we implemented an intensified active data collection through ReMAP focusing on COVID-19-related outcome variables (COVID-19 assessment). Participants had the opportunity to provide weekly feedback on their adaptive capabilities to the pandemic via their smartphone. All participants gave written informed consent. The study was approved by the institutional review board of the medical faculty at the University of Münster and the local data protection officer and conducted according to the guidelines of the declaration of Helsinki.

### 2.2 Study design

Participants of the ReMAP study were recruited from ongoing cohort studies of the Institute of Translational Psychiatry. Details on the respective cohort studies can be found in previous publications (Opel et al., 2019, Richter et al., 2020, Meinert et al., 2019, Redlich et al., 2016, Sindermann et al., 2022). Inclusion criteria for our study were at least two completed COVID-19 assessments via ReMAP; one during lockdown and one during the subsequent period of relaxation. The number of completed assessments per participant ranged from two to twelve. Lockdown and relaxation periods were identified based on a data set by the Leibniz Institute for Psychology Information (ZPID; release 4.0) (Steinmetz et al., 2021; Steinmetz & Batzdorfer, 2020). This data set documents 16 different COVID-related governmental measures of restriction in Germany, with a daily resolution, covering the complete recruitment time of the present study. Based on this data we operationalized a lockdown phase as the period when restrictions in the domains of shopping, school, daycare and gastronomy prevailed (March 21 to April 25, 2020). Restrictions during that time in Germany included a shutdown of all non-essential shops, partial closure of schools and daycare with some exceptions (e.g., for classes in final year of graduation or daycare for children of essential professional groups), and shutdown of gastronomy. This lockdown period was compared to a phase of relaxation during which all the restrictive measures described above were completely suspended (August 17 to November 1, 2020). We identified 151 participants who met the inclusion criteria. 74 participants were diagnosed with MDD and 77 people served as HCs. Presence or absence of MDD or any other lifetime mental disorder was evaluated by means of the Structured Clinical Interview for DSM-IV (Diagnostic and Statistical Manual of Mental Disorders, Fourth Edition) Axis I Disorders (SCID-I) (Segal et al., 2009) as part of the assessment routine and at the time of inclusion in the respective original cohort studies. Using the Childhood Trauma Questionnaire (CTQ; Bernstein et al., 1998) and Walker’s (1999) suggested cut-off values of the five subscales of the CTQ (emotional abuse, emotional neglect, physical abuse, physical neglect and sexual abuse) we identified individuals in both study samples (MDDs and HCs) who experienced maltreatment in their childhood. CTQ data was available for 102 of all 151 participants. According to Walker’s definition, 62 subjects who experienced CM (48 subjects with MDD and 12 HCs) and 40 subjects who did not experience CM (12 subjects with MDD, 28 HCs) participated in the study. Please refer to Table S1 for more information on the study sample.

### 2.3 Data collection

We started data collection on April 6^th^, 2020. The last assessment of the presented study was completed on November 11^th^, 2020. The COVID-19 assessment was conducted once a week and asked participants to reflect on the past seven days. We identified social behavior (direct and indirect contacts) and COVID-19 specific mental health indicators (COVID-19-related fear, feelings of isolation) as outcome variables. We asked about the number of contacts that participants had personally met the day before (direct contacts), and the number of contacts that participants had virtually been in contact with the day before (indirect contacts). COVID-19-related fear was evaluated by a sum-score (0-30) consisting of three separate 10-point scales (0: no fear, 10: great fear) which assessed fear in relation to participants’ own health, fear in relation to the health of close relatives and fear in relation to the pandemics’ economic impact. This set of questions was previously validated by Brosch and colleagues (2022). Lastly, for assessing the feeling of isolation we used the sum score (3-12) of three separate four-point Likert-scales (1: I never feel this way, 4: I often feel this way), which asked about the feeling of being isolated, the feeling of being alone, and the feeling of having no one to turn to. The questions were derived from the revised UCLA loneliness scale (Russell et al., 1980). Participants were instructed that answering all questions was optional and they were free to choose their time of answering whenever items were made available.

### 2.4 Statistical Analyses

Statistics were computed using R statistical software (version 4.3.0; R Core Team).

A two-sided independent *t*-test and a chi-square test were calculated to determine whether subjects with MDD and HCs and subjects with and without the experience of CM differed in age and gender. Moreover, we calculated the mean BDI-II score of the study samples and used a two-sided independent t-test to investigate whether participants with MDD differed from HCs and whether participants with the experience of CM differed from participants without the experience of CM in depressive symptomatology.

Changes in outcome variables between the operationalized lockdown period and the period of relaxation were analyzed using linear mixed models. Linear mixed models allow to examine the independent variables of interest while also taking into account variability within and across participants and items simultaneously (Brown, 2021). Data from all assessments contribute to the comparisons while avoiding an overrepresentation of subjects with the most completed assessments (Newgard & Lewis, 2015). Linear mixed models were a suitable method for our investigations, since a substantial proportion of the variance of the different outcome measures was attributable to within-subject variability rather than between-subject variability (adjusted intraclass correlation coefficients ranging from 0.35 to 0.81). First, we investigated the overall changes in each outcome variable using timepoint (lockdown/relaxation) as the exposure variable. We then added the diagnosis of MDD into each model as a second exposure variable, including an interaction term between timepoint and the diagnosis of MDD, to investigate whether rate of change in the outcome variables over time varied according to the diagnosis of MDD.

We followed a similar strategy when investigating the impact of CM. We explored the overall changes in each outcome variable using timepoint as the exposure variable and then added the experience of CM as an additional exposure variable. Lastly, we investigated the interaction between timepoint (lockdown/relaxation) and the experience of CM when analyzing the overall change in the outcome variables. When evaluating the mixed modeling effects on participants with and without CM we controlled for the effects of the diagnosis of MDD.

For exploratory purposes, whenever we found a significant interaction effect between the diagnosis of MDD or the experience of CM and the change from lockdown to relaxation on the dependent variables, we calculated a three-way interaction, investigating the effect of the interaction between timepoint, the diagnosis of MDD and the experience of CM.

## 3. Results

### 3.1 Study sample

The average age of subjects was 44 (SD 13.76) years. There was no significant age difference, neither between subjects with MDD and HCs (*t*(149)=0.43, *p*=.667) nor between subjects with and without the experience of CM (*t*(100)=0.44, *p*=.657). In total 105 women and 46 men participated in the study. A significant gender difference could neither be found between subjects with MDD and HCs (X*^2^*=1.57, *p*=.210) nor between subjects with and without the experience of CM (*X^2^*=1.21, *p*=.271, Table 1). There was a significant difference in the BDI-II scores between participants with MDD and HCs (*t*(134)=7.68, *p*=˂.001) and between participants with the experience of CM and participants without the experience of CM (*t*(97)=5.22, *p*=˂.001).

**Table 1:**
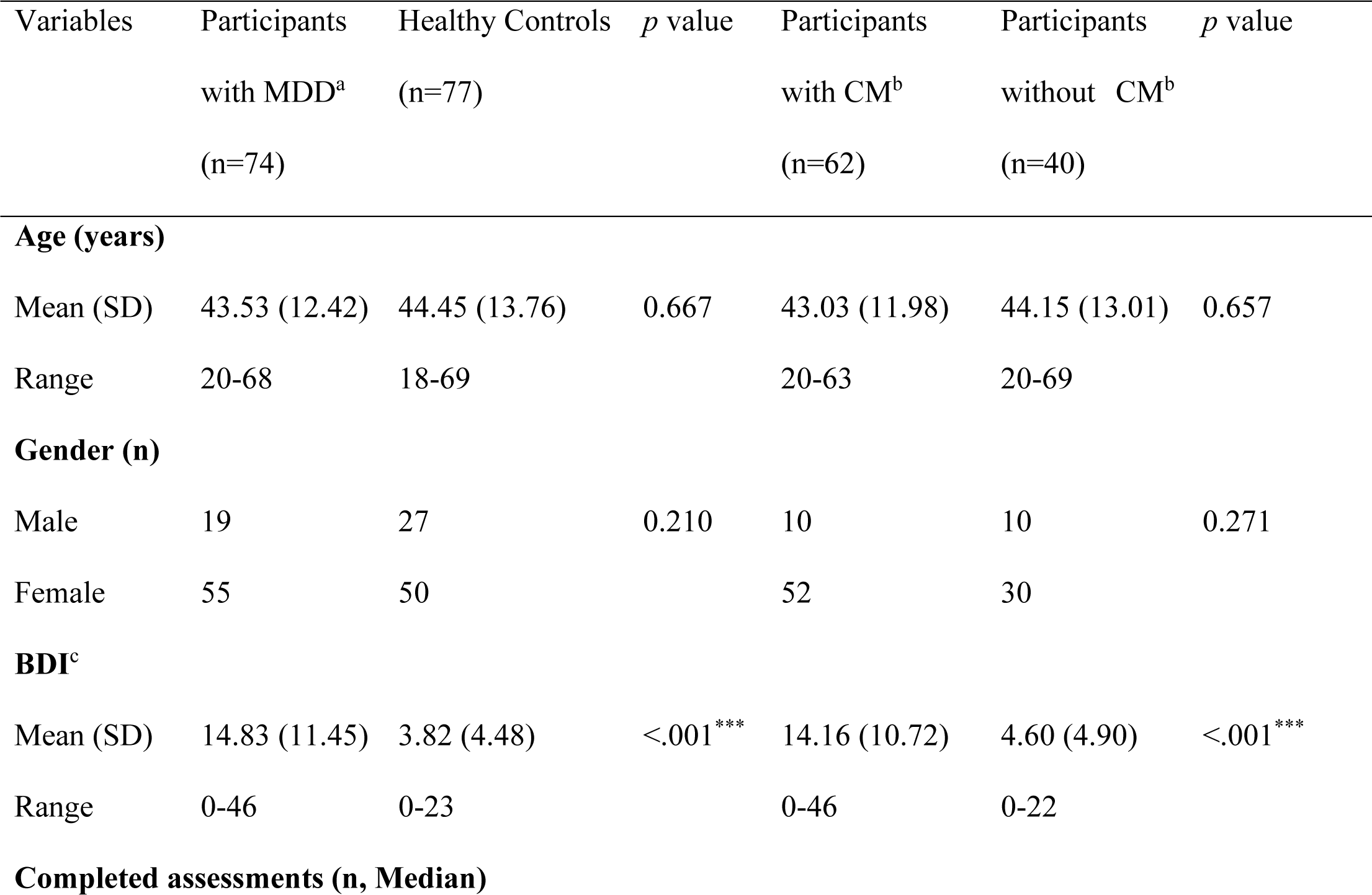
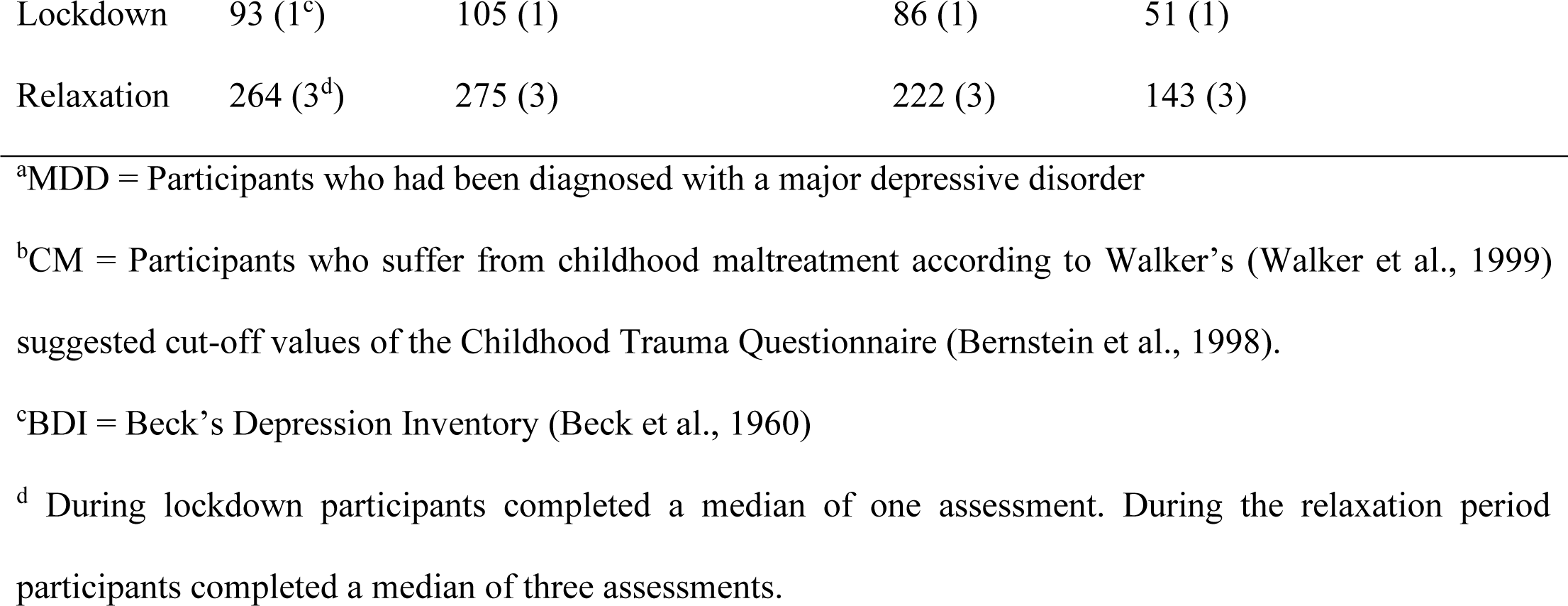
Sociodemographic characteristics and differences between participants with a major depressive disorder (MDD) and healthy controls and between participants who experienced childhood maltreatment (CM) and participants who did not experience CM

### 3.2 Adaptation from lockdown to relaxation

When investigating social behavior in all participants, we found that the number of direct contacts significantly increased (estimated mean score difference:1.52, *CI*:0.88 to 2.16, conditional *R^2^*=0.37, *p*=˂.001), while the number of indirect contacts significantly decreased from lockdown to the relaxation period (estimated mean score difference: -0.91, *CI*:-1.46 to - 0.37, conditional *R^2^*=0.39, *p*=.001, Table 2, Figure 1).

**Figure 1:**
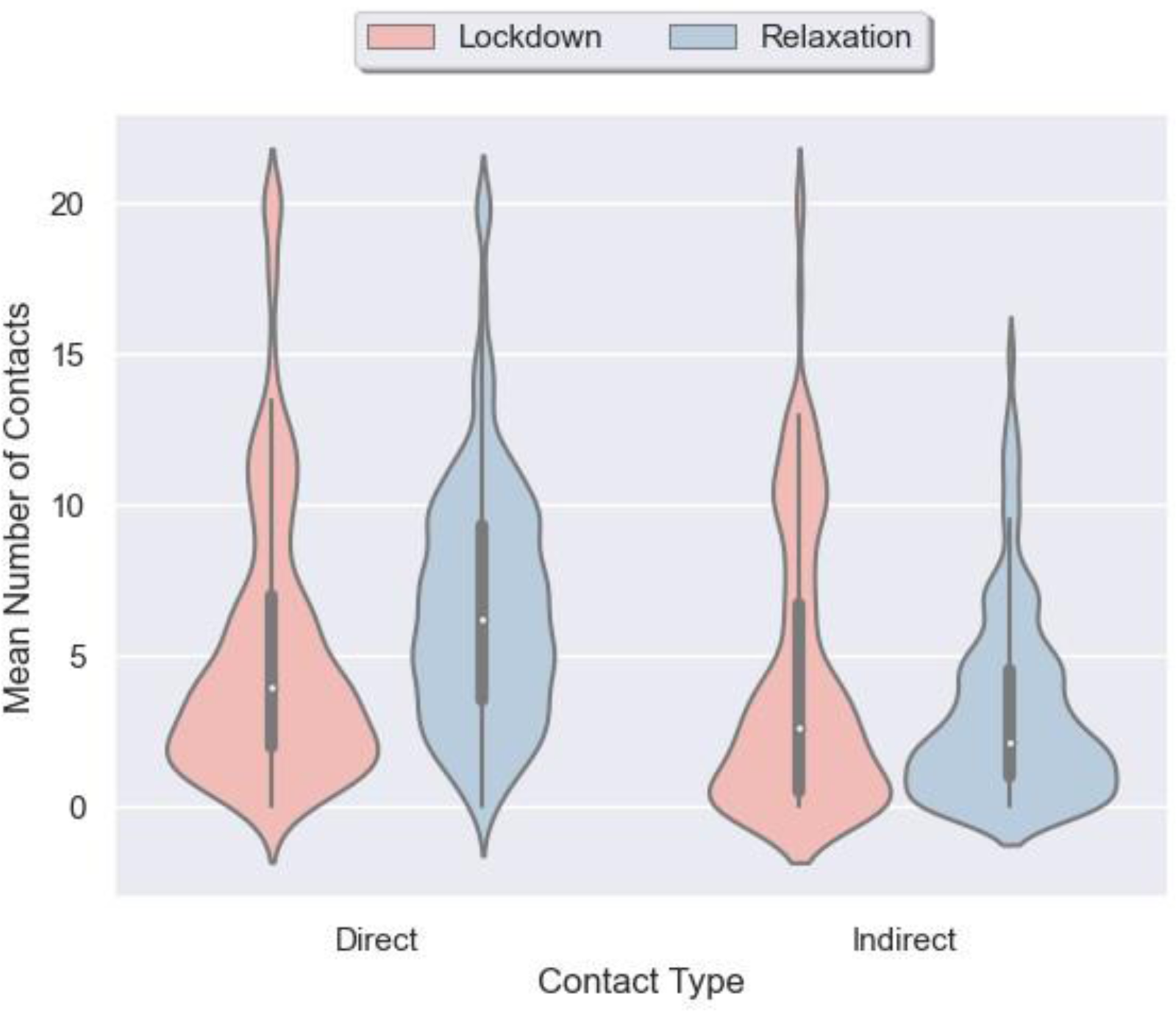
The mean number of contacts of the study sample during the change from lockdown to the relaxation period

**Table 2:**
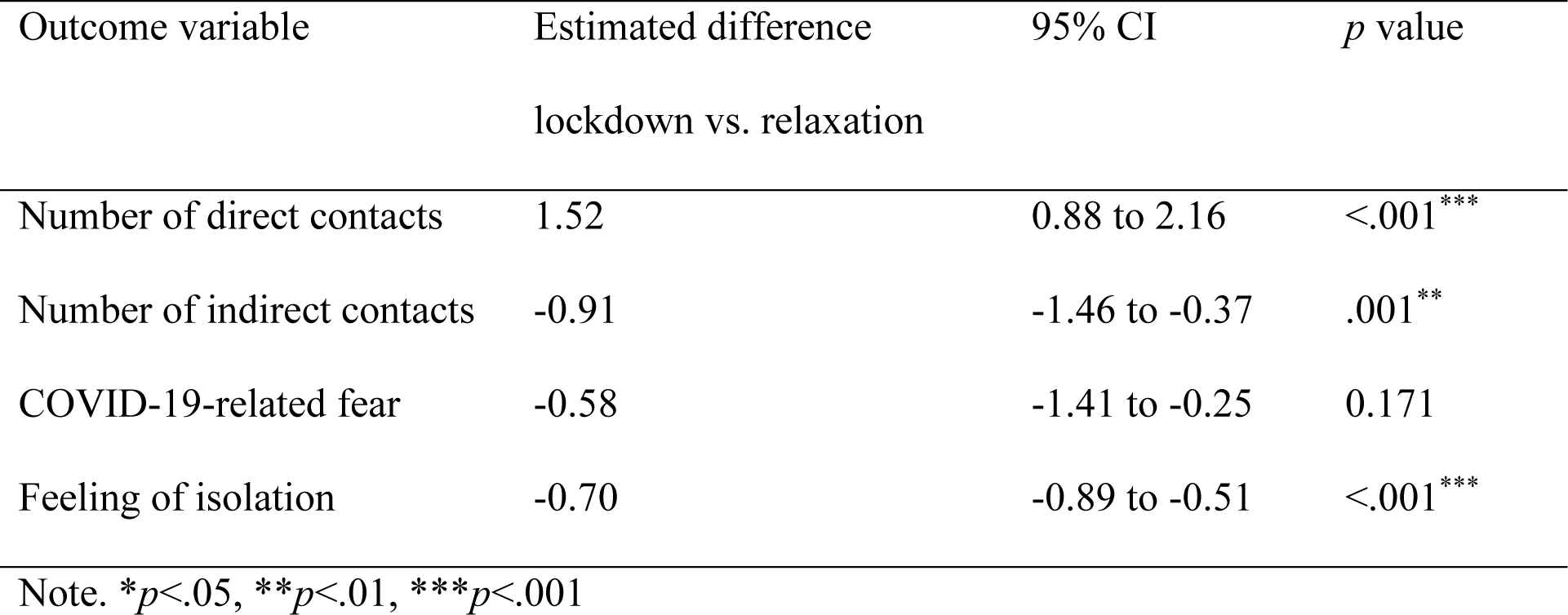
Estimated overall differences in each outcome variable (number of direct contacts, number of indirect contacts, COVID-19-related fear, feeling of isolation) between lockdown and relaxation of the study sample

Regarding the whole study sample COVID-19-related fear (estimated mean score difference: -0.58; *CI*:-1.41 to -0.25, conditional *R^2^*=0.73, *p*=.171) did not change between lockdown and relaxation, while the feeling of being isolated significantly decreased (estimated mean score difference:-0.70; *CI*:-0.89 to -0.51, conditional R*^2^*=0.816, *p*=˂.001; Table 2).

### 3.3 Moderating effect of MDD and CM on adaptation from lockdown to relaxation

We added respective interaction terms between the diagnosis of MDD and the timepoint (lockdown/relaxation) and between the experience of CM and timepoint to examine whether MDD or CM moderated change in outcome variables between lockdown and relaxation (Table 3).

**Table 3:** Changes in all outcome variables (number of direct contacts, number of indirect contacts, COVID 19-related fear, feeling of isolation) that can be attributed to the interaction between the timepoint (lockdown/relaxation) and either the diagnosis of MDD or the experience of CM

|  | Change through the interaction<br>with diagnosis of MDD |  |  | Change through the interaction<br>with the experience of CM |  |  |
| --- | --- | --- | --- | --- | --- | --- |
| Outcome variable | Estimated<br>difference | 95% CI | *p* value | Estimated<br>difference | 95% CI | *p* value |
| Number of direct contacts | 0.03 | -1.25 to 1.32 | 0.961 | 0.39 | -1.23 to 2.02 | 0.641 |
| Number of indirect<br>contacts | 0.43 | -0.66 to 1.52 | 0.441 | 1.38 | -0.04 to 2.82 | 0.058 |
| COVID-19-related fear | 2.28 | 0.64 to 3.92 | 0.007** | 2.80 | 0.79 to 4.82 | 0.007** |
| Feelings of isolation | -0.02 | -0.40 to 0.36 | 0.908 | -0.11 | -0.62 to 0.39 | 0.666 |

Note. \* $p < .05$ , \*\* $p < .01$ , \*\*\* $p < .001$

There was no evidence for an interaction effect of the diagnosis of MDD and timepoint (lockdown/relaxation) on participants’ daily number of contacts, either direct (estimated difference:0.03, CI:-1.25 to 1.32, conditional R*^2^*=0.37, *p*=.961) or indirect contacts (estimated difference:0.43, CI:-0.66 to 1.52, conditional R*^2^*=0.39, *p*=.441). We did not observe a significant interaction effect of CM and timepoint on the number of direct contacts either (estimated mean difference=0.39, CI:-1.23 to 2.02, conditional *R^2^*=0.41, *p*=.641, Figure 2). Even if there was no significant interaction effect between the experience of CM and the change from lockdown to relaxation on the indirect contacts of participants (estimated mean difference=1.38, CI:-0.04 to 2.82, conditional *R^2^*=0.35, *p*=.058), we found that subjects who experienced CM showed no significant change in the number of indirect contacts (estimated mean difference:-0.63, CI:-2.52 to 0.91, *p*=.187), while subjects who did not experience CM showed a significant decrease in their number of indirect contacts (estimated mean difference: -1.98, CI:-3.09 to 0.76, *p*< .001, Figure 2).

**Figure 2:**
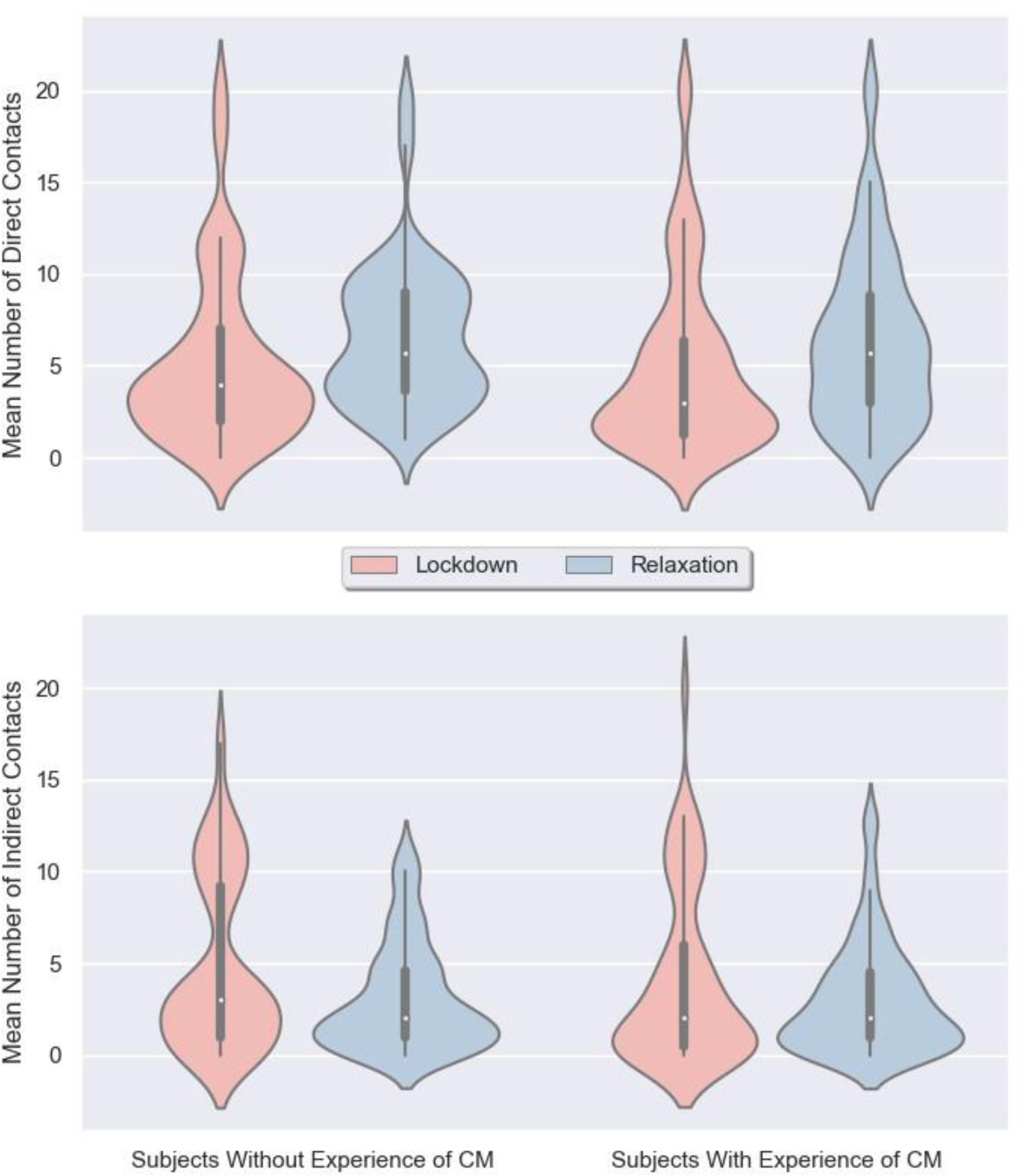
Comparison of the change of direct and indirect contacts of participants who experienced CM and participants who did not experience CM during the change from lockdown to the relaxation period

A significant interaction effect of MDD diagnosis and timepoint (lockdown/relaxation) on the course of COVID-19-related fear (estimated mean difference=2.28, CI:0.64 to 3.92, conditional R*^2^*=0.39, *p*=.007, Figure 3) was observed. HCs showed a decrease in fear (estimated mean difference:-1.65, CI:-2.81 to -0.51, *p*=.005), while fear of participants with a diagnosis of MDD did not change significantly (estimated mean difference:0.63, CI:-0.55 to 1.80, *p*=.298, Table 3). When investigating the interaction effect between the experience of CM and timepoint on COVID-19-related fear a similar phenomenon was identified: The experience of CM moderated an attenuated change of COVID-19-related fear (estimated mean difference=2.80, CI:0.79 to 4.82, conditional R*^2^*=0.76, *p*=.007, Figure 3). Subjects who did not experience CM showed a decrease in COVID-19-related fear (estimated mean difference:-2.31, CI:-3.85 to -0.81, *p*=.003), while the fear of subjects with an experience of CM did not significantly change between lockdown and relaxation (estimated mean difference:0.48, CI:-0.85 to 1.80, *p*=.478, Figure 3).

**Figure 3:**
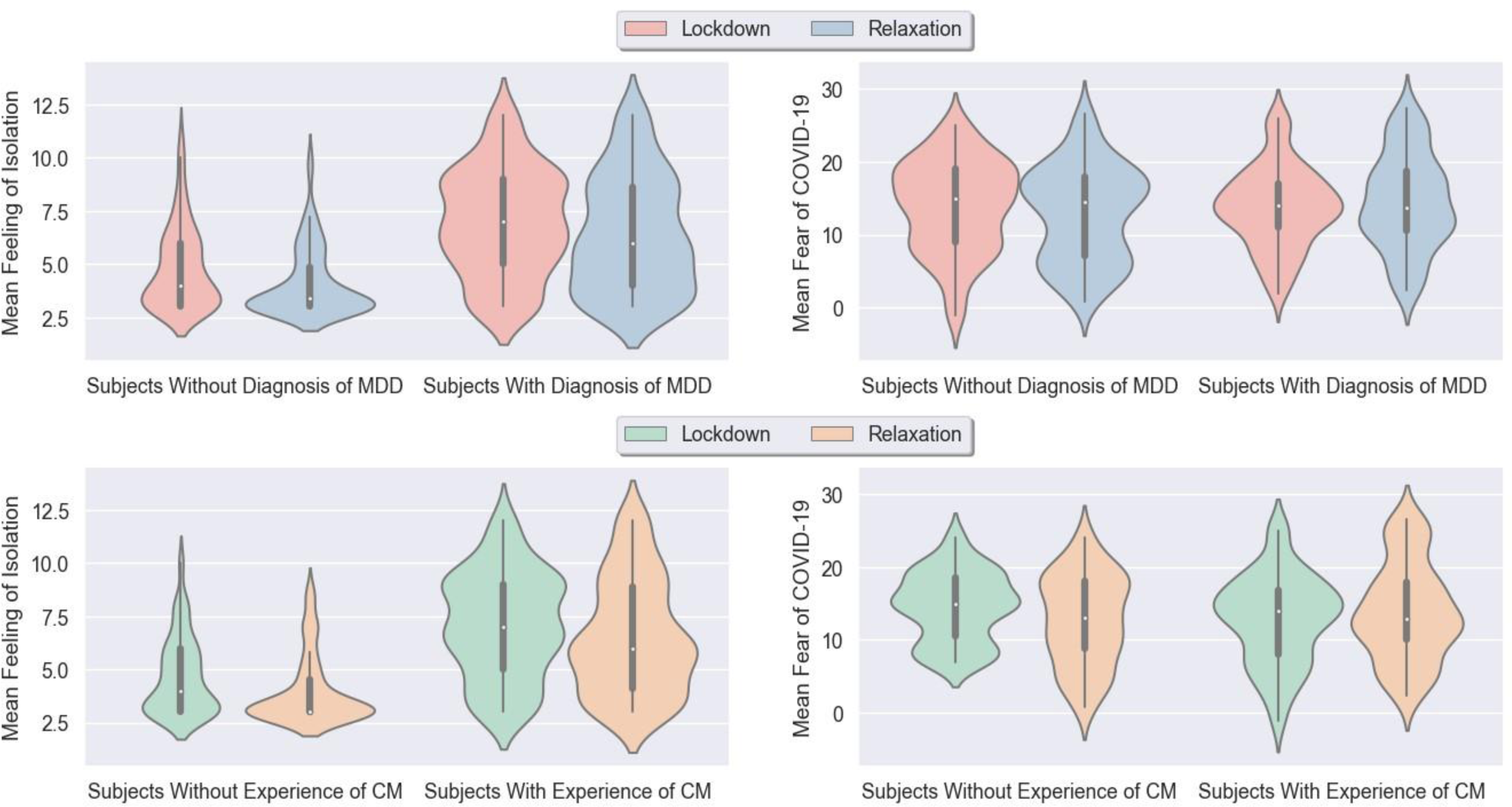
Comparison of the feeling of isolation and COVID-19 related fear in participants diagnosed with MDD and participants without the diagnosis of MDD and in participants who experienced CM and participants without the experience of CM during the change from lockdown to the relaxation period

We did not observe a significant interaction effect between the diagnosis of MDD (estimated difference:-0.02, CI:-0.40 to 0.36, conditional R*^2^*=0.82, *p*=.908, Figure 2) or the experience of CM (estimated mean difference:-0.11, CI:-0.62 to 0.39, conditional *R^2^*=0.81, *p*=.666) and the change from lockdown to relaxation on feelings of isolation.

Additional analyses investigating the effects of the three-way interaction between the change from lockdown to relaxation, the experience of CM and the diagnosis of MDD found no significant association with COVID-19-related fear (estimated mean difference:-2.91, CI:-8.04 to 2.17, conditional R*^2^*=0.76, *p*=.268). However, when exploring the development of COVID-19-related fear of the different subgroups described in Table S1, we found that only participants who were neither diagnosed with MDD nor experienced CM showed a decrease in fear with the change from lockdown to relaxation (estimated mean difference:-2.95, CI:-4.54 to -1.39, *p*< .001). Participants who were either diagnosed with MDD (estimated mean difference:-0.05, CI: -4.07 to 3.90, *p=.*981), experienced CM (estimated mean difference:0.37, CI:-3.45 to 4.31, *p*=.850) or both (estimated mean difference:0.48, CI:-0.93 to 1.89, *p*=.501) showed no significant change in COVID-19-related fear between lockdown and relaxation (Figure S1).

## 4. Discussion

The COVID-19 pandemic has undeniably been a major mental health burden, and for many, it continues to be so. Even as circumstances normalize and the prospect of an upcoming lockdown diminishes, it is likely not the last pandemic most of us will experience in our lifetime. The frequency of emerging infectious diseases is increasing, leading to a rise in the variety of causal diseases (McArthur, 2019; Smith et al., 2014). Consequently, in future pandemic emergencies, it will be crucial to digitally capture social support and mental health status, especially for vulnerable groups and utilize knowledge about mental health impacts of regulations and their relaxation, based on empirical evidence. In this study, we use a smartphone-based data collection for capturing participants’ social cohesion and mental health status and identify participants with MDD and participants with CM as a group particularly vulnerable for sustained perception of fear even in face of allegedly non-critical periods of relaxation.

As restrictions were lifted, participants, in general, became more engaged in direct interpersonal contact again. Additionally, the feeling of isolation decreased with the transition from lockdown to relaxation. However, we made the concerning finding that individuals diagnosed with MDD or those who have experienced CM maintained their fear of COVID-19, even after the pandemic constraints were lifted. Hence, the only subgroup that showed a decrease in fear consisted of participants who neither had an MDD diagnosis nor had experienced CM.

The data suggests that participants with a diagnosis of MDD or a history of CM were able to reconnect with their pre-pandemic social life, although there are indications that subjects who experienced CM held on more to their indirect contacts than the control group. Silveira and colleagues (2022) observed that high levels of social cohesion predict better mental health recovery after lockdown periods. On the other hand, another study showed that low social support was associated with increased suicidality in the first months of relaxation (Farooq et al., 2021). Furthermore, virtual contacts have been found to have a protective influence on psychological well-being during and after the pandemic (Budimir et al., 2021). Social support is closely linked to the feeling of isolation or loneliness, which is one of the most significant public health burdens, leading to an increased risk of mental and physical health problems such as cardiovascular disease, hypertension, diabetes, and infectious diseases (Lancet, 2023). Thus, tracking the development of loneliness throughout lockdown and opening periods is of utmost importance and it is gratifying to see that the feeling of loneliness has decreased, even among vulnerable groups.

The finding that participants who are diagnosed with MDD and who experienced CM maintained their COVID-19 related fear during the change from lockdown to relaxation holds considerable significance. A systematic review and meta-analysis conducted by Alimoradi and colleagues (2022) supports the association between fear of COVID-19 and various mental health factors. Their study revealed that COVID-19-related fear was linked to depression, anxiety, stress and sleeping problems. The association between elevated rates of fear and mental health problems is partly due to a mediating role of COVID-19-related fear in the relationship between intolerance of uncertainty (a characteristic of subjects diagnosed with MDD and those who experienced CM; Hitzler et al., 2022; Pätru et al., 2022) and the onset of depressive symptoms during the pandemic (Voitsidis et al., 2021).

There are limitations to this study. Although we started data collection just 16 days after the start of lockdown, we could not study the immediate effects of the change from pre-pandemic life to lockdown. We also did not have pre-pandemic data to compare our results with. To gain a complete understanding of the impact of COVID-19 and lockdown on individual mental health, it is crucial to compare the outcome variables during relaxation with pre-pandemic data. Future studies that have pre-pandemic data available should consider this. Furthermore, to increase the power of the three-way interaction between timepoint, diagnosis of MDD, and experience of CM, a larger sample size with larger subgroups is needed. The results of the interaction presented in this study can only be considered as tentative evidence. Lastly, we want to address that the effect of the pandemic on the mental health of the general population is still a major topic of discussion (Ahmed et al., 2023). While there are also studies that identify improvements in mental health of the general population in Germany during the pandemic, these same studies call attention to vulnerable subgroups that suffer particularly from the alternating cycles of lockdown and relaxation and should therefore receive special attention in research (Ahmed et al., 2023; Ahrens et al., 2021a, 2021b).

Our study results suggest that the psychological impact of COVID-19-related measures might be sustained in these vulnerable groups. The experience of CM and previous MDD diagnoses seem to impair adjustment processes regarding COVID-19-related fear. Subgroups may be at increased risk for further mental health problems in the post-pandemic phase of COVID-19 and future pandemic outbreaks. Thus, our findings highlight the need for public health considerations, psychosocial care and prevention efforts targeting mental health in highly vulnerable subgroups such as individuals who are diagnosed with MDD or who experienced CM. In this regard, management of anxiety and improving reintegration into personal social life need to be addressed in therapy and social discussions with regard to vulnerable subgroups in the post-pandemic phase.

## Data Availability

All data produced in the present study are available upon reasonable request to the authors

## Notes

### Competing Interest Statement

The authors have declared no competing interest.

### Funding Statement

Funding was provided by the Interdisciplinary Center for Clinical Research (IZKF) of the medical faculty of Muenster (Grant SEED 11/19 to NO), as well as the ''Innovative Medizinische Forschung'' (IMF) of the medical faculty of Muenster (Grants OP121710 to NO).

### Author Declarations

Institutional review board of the Medical Faculty, University of Muenster, Germany gave ethical approval for this work.

