## Supplementary material for "The impact of depression and childhood maltreatment experiences on psychological adaptation from lockdown to relaxation periods during the COVID-19 pandemic": Table S1

**Table S1:** The distribution of the experience of childhood maltreatment (CM) in the study sample

| Maltreatment status | Participants with a major depression disorder (N=74) | Healthy controls (N=77) |
|---|---|---|
| History of CM (N=62) | 48 | 14 |
| No history of CM (N=40) | 12 | 28 |

We performed the childhood Trauma questionnaire (CTQ) (Bernstein et al., 2003) in 102 of all 151 participants.

CM was assessed according to Walker's suggested cut-off values of the CTQ (Walker et al., 1999).

***References***

Bernstein, D. P., Stein, J. A., Newcomb, M. D., Walker, E., Pogge, D., Ahluvalia, T., Stokes, J., Handelsman, L., Medrano, M., Desmond, D., & Zule, W. (2003). Development and validation of a brief screening version of the Childhood Trauma Questionnaire. *Child Abuse and Neglect*, *27*(2), 169–190. https://doi.org/10.1016/S0145-2134(02)00541-0
