## Supplementary material for "The impact of depression and childhood maltreatment experiences on psychological adaptation from lockdown to relaxation periods during the COVID-19 pandemic": Figure S1

**Figure S1:** Comparison of COVID-19 related fear in participants who are not diagnosed with MDD and who did not experience CM, participants who are diagnosed with MDD and who did not experience CM, participants who experienced CM but are not diagnosed with MDD and participants who are diagnosed with MDD and who experienced CM during the change from lockdown to the relaxation period

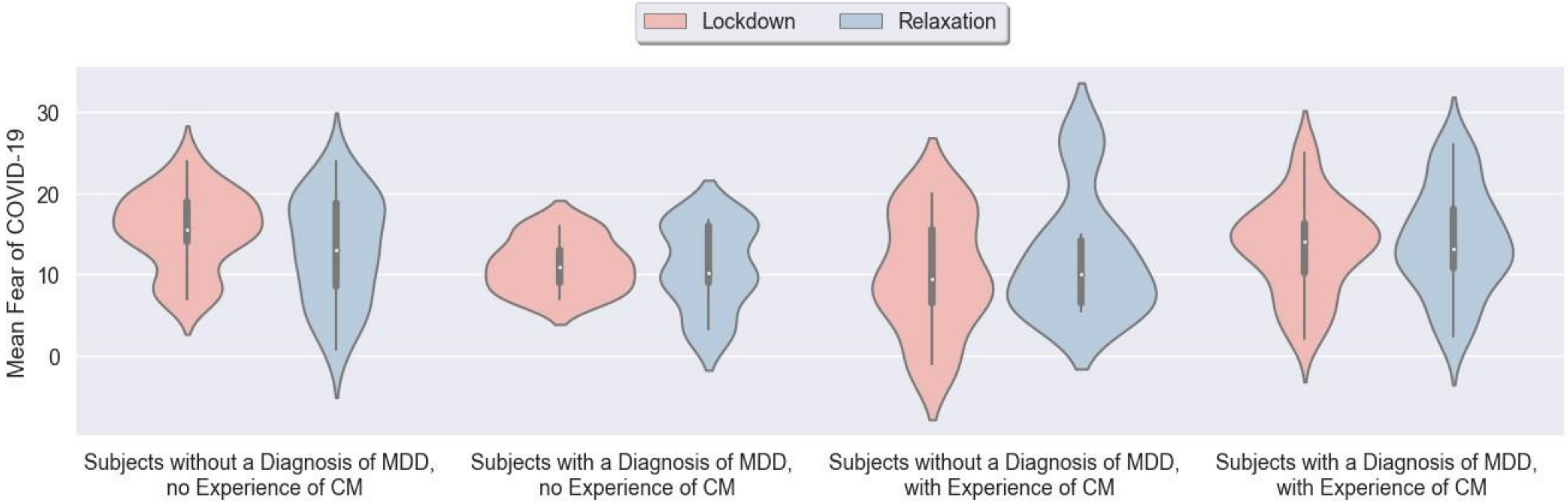
